# Do Rule-Derived Labels Improve Prediction When Reported Labels Are Limited? A Two-Site Machine Learning Study

**DOI:** 10.64898/2026.11.12.26367890

**Authors:** Talim Hossain Sadi, Chowdhury Aseer Ruthbah

**Affiliations:** Department of Computer Science and Engineering, BRAC University, Dhaka, Bangladesh

**Keywords:** weak supervision, limited-label learning, label budget, probability calibration, class imbalance, antimicrobial resistance

## Abstract

**Background:** Rule-derived (weak) labels can enlarge a training set when reported labels are scarce. Their value depends on label quality, on how the extra records shift the training distribution, and on whether the comparison accounts for reported labels set aside for tuning or calibration.

**Methods:** We ran an exploratory label-budget experiment on *Escherichia coli* urine cultures from two de-identified electronic health record datasets: Stanford ARMD and UT Southwestern ARMD-UTSW. The task was to predict reported non-susceptibility to cefepime, cefoxitin and ceftazidime (Stanford) and to cefepime and ceftazidime (UT Southwestern). Reported-label budgets of 10%, 25%, 50% and 100% of development patients were drawn over five patient-disjoint outer splits. Logistic regression and histogram gradient boosting were trained with reported labels only, or with added rule-derived labels using equal weights, a tuning-selected weight, class-proportion adjustment or prior-probability correction. These were compared with an all-budget reported-only baseline that used every budgeted reported label for training. Stanford used the inferred labels released with the dataset and a feature set including resistance history. UT Southwestern used researcher-derived labels from Stanford rule text, which were not validated for that site, and an age-only primary model. Brier-score differences were assessed with paired patient-cluster bootstrap intervals.

**Results:** All rule-derived labels were susceptible. Equal-weight augmentation lowered mean predicted risk well below observed prevalence (for example, Stanford cefepime at the 10% budget: 0.35% versus 4.85%) and had a higher Brier score than the all-budget baseline in every drug, model and budget summary. The tuning-selected weight did not consistently improve on the all-budget baseline. Its median Brier difference was positive in 38 of 40 drug, model and budget summaries. Its paired intervals lay above zero in 100 of 296 Stanford analyses and 31 of 198 UT Southwestern analyses, and below zero in 3 and 5. The tuning set often selected a weight of zero (146 of 296 and 153 of 198 analyses). At UT Southwestern, the age-only models showed little discrimination (median AUROC 0.49 to 0.54), so those comparisons mainly reflect calibration.

**Conclusions:** Under these designs, rule-derived labels did not consistently improve probability prediction over a reported-only model that used the whole reported-label budget for training. Evaluations of weak supervision should include such a baseline and should report calibration as well as discrimination.

## 1. Introduction

Supervised models need labelled examples, and labels are often the expensive part of a machine learning project. Weak supervision replaces part of the manual labelling with cheaper sources such as heuristic rules, knowledge bases or other imperfect labellers. Snorkel made this approach practical by letting users write labelling functions and combining their noisy outputs without ground-truth labels [1]. Since then, many methods for aggregating and using weak labels have been proposed. The WRENCH benchmark was built because these methods had been evaluated on different, often private, datasets with different weak sources and evaluation protocols, which made results hard to compare [2]. Later work found that relatively simple designs can match more complex weak-supervision pipelines on standard benchmarks [3].

How the evaluation counts labelled data matters. Zhu et al. showed in natural language processing tasks that many weakly supervised methods depended on clean labelled validation data. Training directly on those clean examples largely removed the advantage of the more sophisticated methods [4]. Methods that learn example weights from a small clean validation set, such as learning-to-reweight [5], have the same property: the validation labels are part of the method’s labelling cost. Our study asks a related question in a different setting. When a fixed number of reported labels is available, is it better to keep some for choosing how rule-derived labels are used, or to train a reported-only model on all of them? Zhu et al.’s findings motivate this comparison; they do not show what will happen in tabular clinical microbiology data.

Weak labels are also not uniformly useful. In an empirical study of generative models trained with weak supervision, Hernández-González and Pérez found that weakly labelled data helped when fully labelled data were limited, and that the benefit depended on how much supervision information the weak labels carried and how informative it was [6]. Rule-derived labels in our setting carry a particular kind of information: the rules we study only ever imply susceptibility, so every added label belongs to the majority class.

Adding labels from one class changes the class balance of the training data, and class balance affects predicted probabilities. In clinical risk prediction, corrections for class imbalance such as undersampling, oversampling and SMOTE did not improve discrimination and produced badly miscalibrated risks [7]. Analyses of random resampling reached similar conclusions and showed that the resulting miscalibration could largely be reversed by a correction for the changed class prior [8]. Calibration is therefore a central part of evaluating probability predictions, not a secondary check [9]. When two data sources differ only in class proportions, the label-shift assumption holds: the class-conditional feature distribution is the same and only the class marginal changes [10]. Under that assumption, predicted probabilities can be adjusted to a new prior with a simple closed-form formula [11]. Rule-derived labels need not satisfy this assumption, because they are attached to cultures that were selected differently from those with reported results.

Antimicrobial susceptibility data provide a natural test case. The Stanford Antibiotic Resistance Microbiology Dataset (ARMD) releases reported susceptibility results and, separately, implied susceptibility labels inferred by linking results between related antibiotics according to Stanford microbiology laboratory protocols based on CLSI standards [12]. The UT Southwestern dataset (ARMD-UTSW) has a similar structure but no separate inferred-label file [13]. In UT Southwestern data, weak labels therefore have to be constructed by the researcher, and their validity for that laboratory cannot be assumed.

We compare reported-only training with several ways of adding rule-derived labels at fixed reported-label budgets, at both sites. The main comparator is an all-budget reported-only baseline that spends the whole budget on training. This is an empirical comparison of established techniques, not a new algorithm. The UT Southwestern analysis refits the models on UT Southwestern data with a different feature set and differently constructed labels. It is therefore an exploratory independent-site replication of the design, not an external validation of a fixed Stanford-trained model in the sense used in prediction-model research [14]. We used the TRIPOD+AI statement [15] as a guide for reporting the prediction-model components.

## 2. Methods

### 2.1 Data and prediction task

We used two publicly released, de-identified microbiology datasets derived from electronic health records: Stanford ARMD (dataset release of 22 October 2025) [12] and ARMD-UTSW (Dryad version of 26 September 2025) [13]. From each, we used the culture cohort, demographic and prior-resistance files. At Stanford we also used the released implied-susceptibility file.

The unit of analysis was a culture order, identified by the composite key of patient, encounter and order identifiers, all read as strings. We selected *E. coli* urine cultures with a complete key. At UT Southwestern, culture orders linked to more than one patient or encounter would have been excluded; none were found. The outcome for each target antibiotic was reported non-susceptibility: Resistant or Intermediate = 1, Susceptible = 0. Other reported values (for example, inconclusive) were ignored, and a culture whose valid reported results disagreed on the binary outcome was excluded. The organism is treated as known, so the task is not prediction of the organism before culture.

Target antibiotics were cefepime, cefoxitin and ceftazidime at Stanford, and cefepime and ceftazidime in the UT Southwestern modelling experiment. UT Southwestern cefoxitin was audited but not modelled: it had 56 reported cultures with 3 non-susceptible results, below the prespecified minimum of 50 cultures per class.

### 2.2 Rule-derived labels

#### Stanford

We used the released implied-susceptibility labels as provided, applying the same binary mapping. Keys with conflicting implied labels, keys that did not match an *E. coli* urine culture, and keys that also had a reported result for the target were excluded. All released implied labels for these targets were susceptible.

#### UT Southwestern

We applied the *E. coli* rules from the Stanford rule file as research weak-labelling heuristics. Cefepime and ceftazidime: susceptible if susceptible to ceftriaxone, cefotaxime or cefazolin. Cefoxitin: susceptible if susceptible to cefazolin. These are Stanford laboratory reporting rules. They were not independently validated for UT Southwestern, and we do not present them as UT Southwestern labels or as rules suitable for patient care. We used a conservative version fixed before training. A culture received a weak susceptible label only if at least one source antibiotic was reported susceptible, no source antibiotic was reported non-susceptible, and no source antibiotic had conflicting results on that culture.

Cultures with any reported result for the target received no weak label. Missing results never produced a label. As a diagnostic, rule predictions were compared with reported target results on cultures that had both. These cultures were selected for testing of both drugs, so agreement on them may not carry over to the cultures that receive weak labels.

While preparing the UT Southwestern analysis, we checked whether the rule text reproduces Stanford’s released labels. Applying the cefoxitin rule (susceptible to cefazolin, no reported cefoxitin result) to Stanford *E. coli* urine cultures gave 24,217 cultures. Of these, 18,123 had a released Stanford implied label and 6,094 did not. The released labels therefore reflect conditions beyond the rule text, and labels derived from the same text at UT Southwestern are not equivalent to them.

### 2.3 Predictors

#### Stanford primary features

age band; gender code (an unordered category; no meaning is given to the codes); ordering mode; and, for any resistance, resistance to the target antibiotic and *E. coli* resistance, the number of prior-resistance records, the smallest day offset and the number of records within 30, 90 and 365 days (log1p-transformed; no history encoded as 0 records and 99,999 days). Positive values of ‘resistant_time_to_culturetim’ were read as days before the culture, and zero, negative and non-numeric offsets were excluded. Neither the direction of this offset nor when earlier results became available is independently verified.

#### UT Southwestern primary features

age band only. Gender was excluded because its coding is not known to match Stanford’s. The UT Southwestern documentation does not describe an ordering-mode field, and it describes the history offset only as days between the culture order and the susceptibility result, which does not establish its direction or availability. A reported-only sensitivity analysis added history features under the Stanford assumption; it is reported in the exported results but not used for the main comparisons. Because the feature sets differ, absolute performance is not comparable between sites. Identifiers, the current result, label source, rule provenance and same-culture results for other antibiotics were never predictors.

### 2.4 Patient allocation and reported-label budgets

Figure 1 summarises the design. For each of five fixed salts, each patient was assigned by SHA-256(salt + patient identifier), using the first eight hexadecimal digits modulo 100. Values of 80 to 99 formed the outer test set; values of 0 to 79 formed the development set, with a training role (0 to 59), tuning role (60 to 69) or calibration role (70 to 79). A patient’s role does not depend on the budget, and all cultures of a patient stay in that patient’s role. Development patients with at least one reported culture for the drug were ordered by a second salted hash for each budget sample. A budget fraction of 10%, 25%, 50% or 100% took a prefix of this order, so budgets are nested within a sample. Three budget samples were drawn for each fraction below 100%, and one at 100%, giving 15 planned analyses per fraction below 100% and 5 at 100% for each drug and model. Rule-derived labels came only from development patients with the training role and were not limited by the budget. Tuning, calibration and test patients never contributed rule-derived records. Every method within an allocation used the same patients, and the outer test set was never resampled. The same salts and cut points were applied at both sites; because the patients differ, the partitions themselves are site-specific.

**Figure 1.**
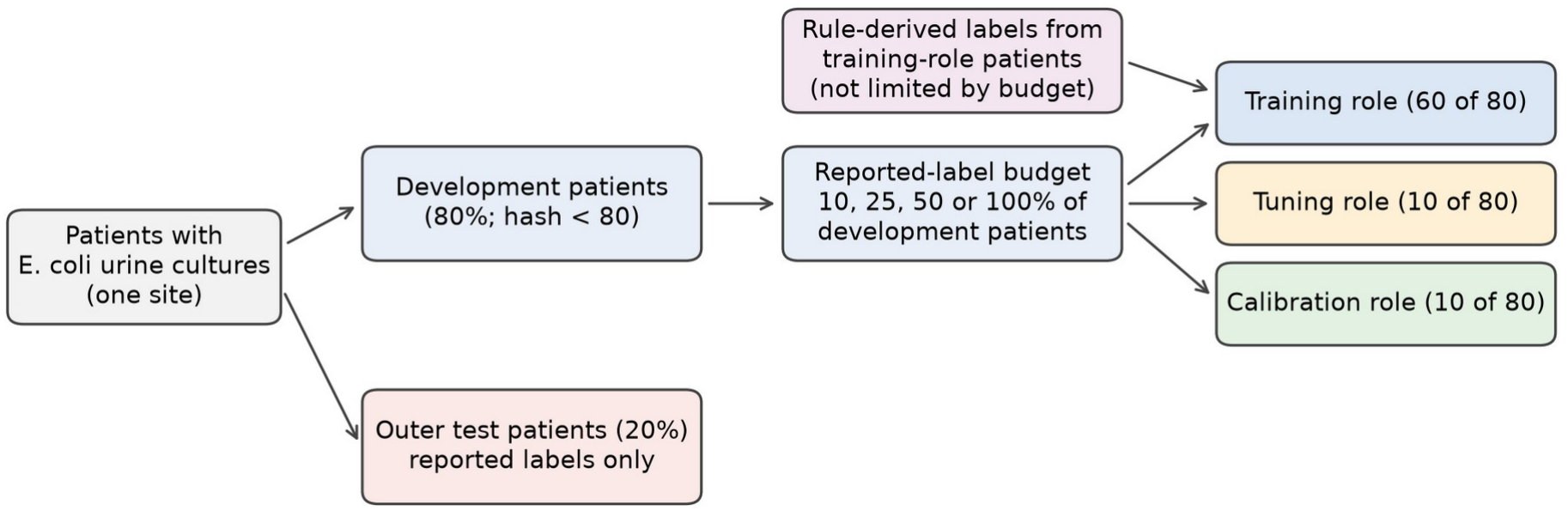
Design of one allocation. Within an allocation, every method uses the same patients, and test patients are never used for fitting, preprocessing, weight selection or recalibration. The all-budget baseline trains one model on the budgeted training, tuning and calibration reported labels together.

### 2.5 Models and compared methods

Both model classes used fixed settings without tuning: L2-penalised logistic regression (C = 1, up to 1,000 iterations) and histogram gradient boosting (100 iterations, 7 leaves, at least 40 samples per leaf, L2 regularisation 5, no early stopping), as implemented in scikit-learn 1.8.0 [16]. One-hot encoding and standardisation were fitted on the training reported cultures of each allocation. Sample weights were rescaled to a mean of 1. Table 2 lists which resources each method uses. Let π_R be the non-susceptible proportion among reported training cultures and π_M the proportion after adding rule-derived cultures.

The selected-weight and recalibrated methods use part of the budget for tuning or calibration, so they train on fewer reported labels than the all-budget baseline. The all-budget comparison therefore measures the combined effect of adding rule-derived labels and of holding reported labels out for selection. Recalibrated methods were also compared with the recalibrated reported-only model, so that both sides had the same calibration resources. An analysis was unavailable if the training or test set had fewer than 10 positive or 10 negative cultures. Weight selection required at least 5 of each class in tuning, and recalibration at least 10 of each in calibration.

### 2.6 Evaluation

On the outer test cultures we computed AUROC; average precision for the non-susceptible class (the AUPRC summary used here); Brier score [18]; log loss; observed prevalence; and mean predicted risk. We also computed calibration-in-the-large (the intercept of a logistic model with logit p as offset; 0 ideal) and calibration slope (the coefficient of logit p; 1 ideal) [9]. Both calibration measures were fitted by unpenalised maximum likelihood and reported as missing when the fit did not converge or diverged. Training and prediction times were recorded. Stanford results were computed on a 2-CPU Linux container and UT Southwestern results in Google Colab, so runtimes are not comparable between sites.

For Brier-score differences we used a paired patient-cluster bootstrap [19] with 2,000 resamples. For each drug and outer split, one set of patient resamples was drawn and shared by every budget, sample, model and method. All cultures of a drawn patient entered with that patient’s multiplicity, and the 2.5th and 97.5th percentiles of the replicate differences formed the interval. A positive difference means the compared method has a worse Brier score than its baseline. These intervals describe uncertainty from test patients only. They condition on the fitted models and do not include variation from budget sampling, training patients or refitting. That variation is summarised separately across outer splits (median and range of split means) and budget samples. The outer splits share patients, and the data had been explored in earlier analyses, so the splits are repeated exploratory evaluations, not independent replications. Many comparisons were made without multiplicity adjustment.

Analyses were run with Python and pinned versions of numpy (2.3.5), pandas (2.2.3) and scikit-learn (1.8.0). The Stanford results reported here come from the same notebook run on the same files as an earlier Colab run. For the methods shared by both runs, test Brier scores agreed within 1.3 × 10^−8^ and AUROC within 3.1 × 10^−5^.

## 3. Results

### 3.1 Cohorts and label budgets

Table 1 summarises the cohorts. At Stanford, 16,197 cefepime, 29,973 cefoxitin and 37,222 ceftazidime reported cultures were included, with non-susceptibility of 4.5%, 11.8% and 7.9%. Stanford released 36,420, 18,175 and 16,327 implied labels for these drugs, all susceptible. At UT Southwestern, 19,759 cefepime cultures (1,208 non-susceptible; 6.1%) and 3,478 ceftazidime cultures (1,321 non-susceptible; 38.0%) were included. The high ceftazidime prevalence is consistent with selective testing of that drug, but we did not investigate why it was tested. The conservative heuristics produced 13,920 cefepime and 29,066 ceftazidime weak labels on cultures without a reported target result. Only those from training-role patients entered training: in the first outer split, 8,423 and 17,535. UT Southwestern ceftazidime test sets were small: in the first outer split, 711 cultures with 306 positives. Supplementary Table S1 gives the budget allocation. At the 10% budget, the UT Southwestern ceftazidime budget contained a median of 274 reported cultures, with 31 tuning and 31 calibration cultures.

**Table 1.**
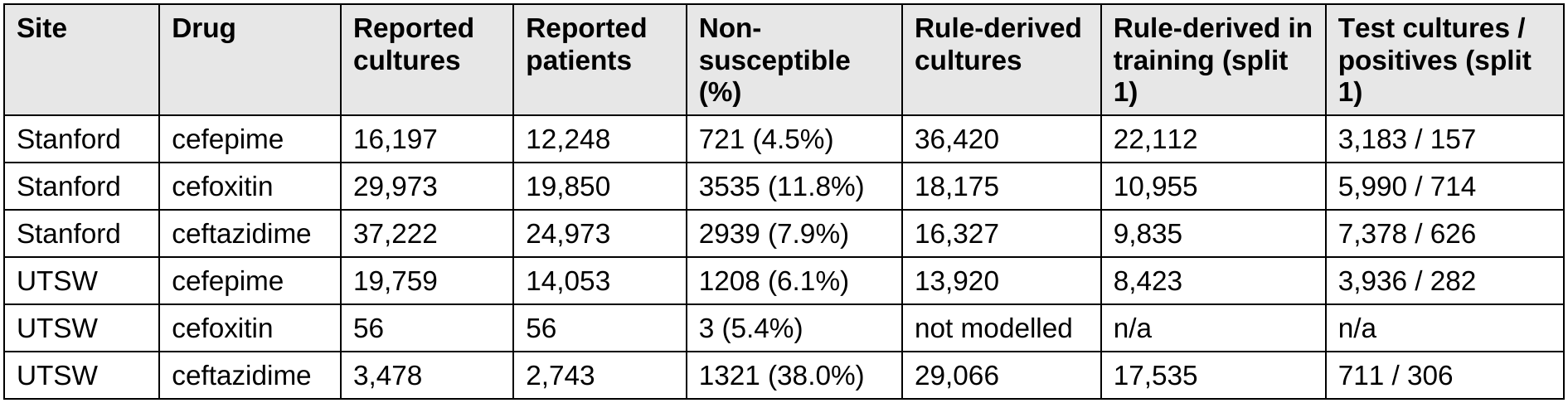
Analysis cohorts. Rule-derived cultures: released implied labels (Stanford) or conservative research heuristics (UT Southwestern), excluding cultures with a reported target result. Training rule-derived cultures and test counts are for the first outer split.

**Table 2.** Compared methods. π_R and π_M are the non-susceptible proportions in reported and in mixed (reported plus rule-derived) training data.

| Method | Definition |
|---|---|
| Reported only | Budgeted training-role reported cultures, weight 1. |
| Equal weight | Reported plus all eligible rule-derived training cultures, weight 1. |
| Selected weight | Rule-derived cultures weighted α ∈ {0, 0.05, 0.1, 0.25, 0.5, 1}, chosen by Brier score on the budgeted tuning cultures (ties to the smaller α). α = 0 is the reported-only model. |
| Class-adjusted | Mixed training data with class weights π_R/π_M and (1 − π_R)/(1 − π_M), which restore the reported class proportions. |
| Prior-corrected | Equal-weight model with predicted odds multiplied by [π_R/(1 − π_R)]/[π_M/(1 − π_M)] [11]. Valid only under pure label shift; it is a known-prior adjustment, not the black-box shift estimator of Lipton et al. [10]. |
| Recalibrated reported only / selected weight | Sigmoid recalibration (logit p̃ = a + b·logit p) [17] fitted by maximum likelihood on the budgeted calibration cultures; run only with at least 10 positive and 10 negative calibration cultures. |
| All-budget reported only | One model trained on all budgeted reported cultures (training, tuning and calibration roles together); no rule-derived labels, no tuning, no recalibration. |

### 3.2 Rule diagnostics at UT Southwestern

Table 3 summarises the UT Southwestern rule diagnostics. Applying the rule text as written, rule predictions disagreed with 20 of 18,220 co-tested reported cefepime results and 75 of 2,210 ceftazidime results. Under the conservative handling, which excluded 2,014 cultures where source antibiotics disagreed and 8 with conflicting source results, the figures were 16 of 16,690 and 17 of 1,553. These counts describe a selected co-tested subset and are not error rates for the weak-label training pool.

**Table 3.** UT Southwestern rule diagnostics. Disagreement = rule predicts susceptible, reported result non-susceptible, among cultures with both.

| Target | Cultures with a susceptible source | Excluded: sources disagree | Excluded: conflicting source | Disagreement, rule text | Disagreement, conservative | Weak labels without reported target |
|---|---|---|---|---|---|---|
| cefepime | 32,641 | 2,014 | 8 | 20 / 18,220 | 16 / 16,690 | 13,920 |
| ceftazidime | 32,641 | 2,014 | 8 | 75 / 2,210 | 17 / 1,553 | 29,066 |
| cefoxitin | 24,368 | 0 | 0 | 2 / 50 | 2 / 50 | 24,318 |

### 3.3 Mean predicted risk

Adding all rule-derived labels with equal weight lowered mean predicted risk well below the observed prevalence at both sites (Figure 2), most strongly at small budgets where rule-derived records dominated the training data. For logistic regression at the 10% budget at Stanford, median mean predicted risk fell from 4.47% (reported only) to 0.35% (equal weight) for cefepime, from 12.21% to 2.88% for cefoxitin, and from 7.93% to 2.60% for ceftazidime. Observed test prevalences were 4.85%, 11.92% and 8.12%. At UT Southwestern the shift was larger for ceftazidime, from 35.0% to 0.42% against an observed 37.4%, because about 17,500 susceptible weak labels were added to a budget of a few hundred reported cultures. The selected weight, class adjustment and prior correction largely removed this shift, and the all-budget baseline stayed close to the observed prevalence.

**Figure 2.**
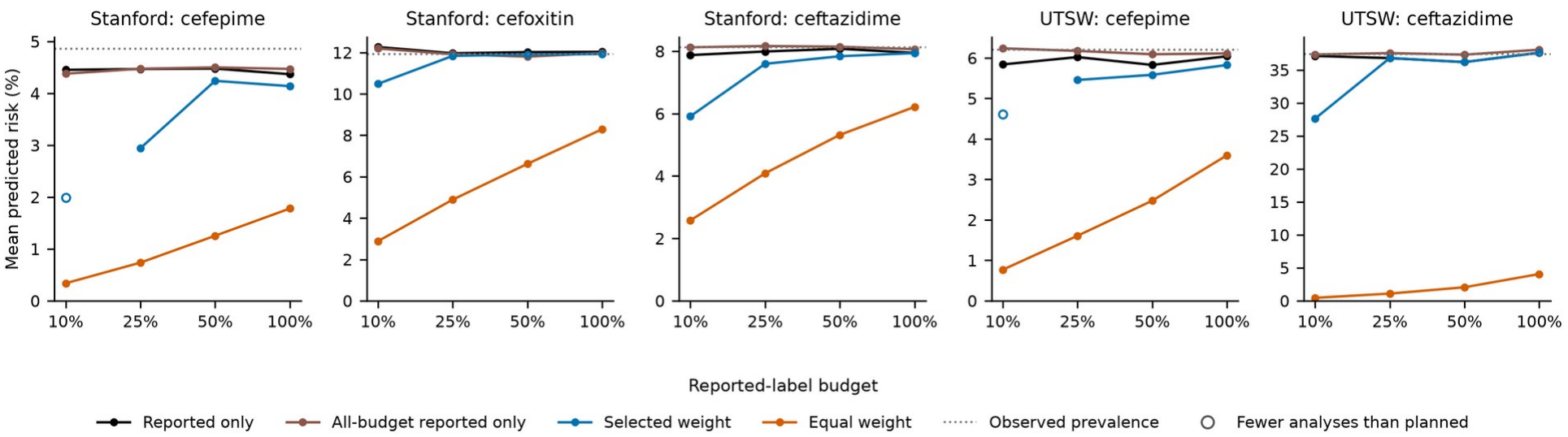
Mean predicted risk on the outer test set by reported-label budget (logistic regression). Each point is the median across outer splits of the split mean over budget samples. Hollow points rest on fewer analyses than planned and are not joined to neighbouring points. Dotted line: observed prevalence (median across splits).

### 3.4 Comparison with the all-budget reported-only baseline

Table 4 and Figure 3 summarise Brier-score differences against the all-budget baseline, and Supplementary Table S2 gives the test metrics. Equal-weight augmentation had a higher median Brier score than the all-budget baseline in every drug, model and budget summary at both sites. Its intervals lay above zero in 227 of 300 Stanford analyses and 191 of 200 UT Southwestern analyses, and never below zero. At UT Southwestern ceftazidime the difference was large: a median of +0.127 at 10% for logistic regression, against a baseline Brier score of about 0.24.

**Figure 3.**
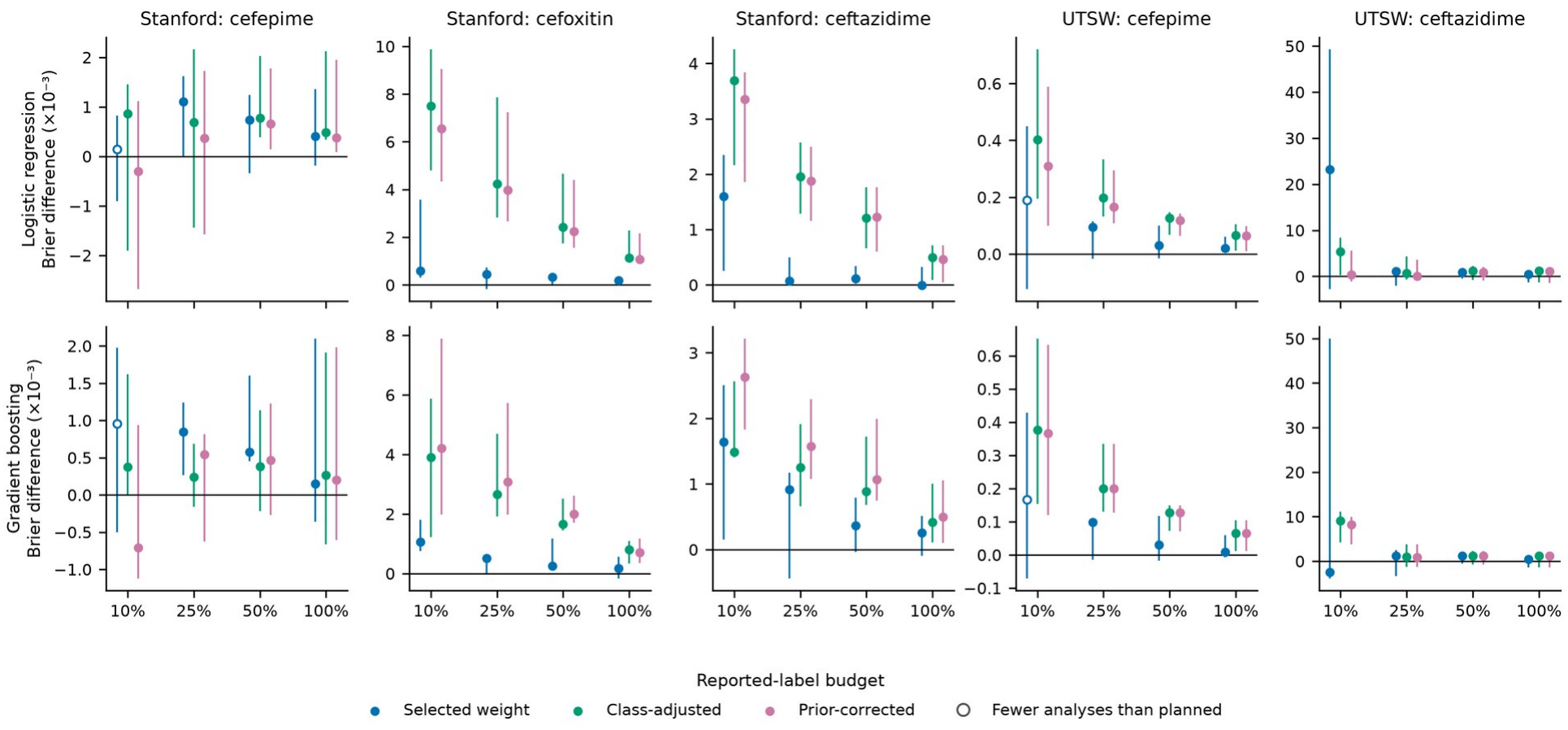
Brier-score difference (× 10^−3^) between each rule-derived method and the all-budget reported-only baseline; positive values mean the rule-derived method is worse. Point: median across outer splits of the split mean over budget samples; bar: range across outer splits. Hollow points rest on fewer analyses than planned. Equal-weight results are in Table 4; their scale would hide the other methods. Note the different y-axis scales.

**Table 4.** Brier-score difference between rule-derived methods and the all-budget reported-only baseline, × 10^−3^ (positive = rule-derived method worse). Each cell: median across analyses (minimum to maximum); number of 95% paired bootstrap intervals above zero / below zero / including zero. Planned analyses: 15 at 10%, 5 at 100%; fewer are noted. LR, logistic regression; HGB, histogram gradient boosting.

| Site | Drug | Model | Method | Budget 10% | Budget 100% |
|---|---|---|---|---|---|
| Stanford | cefepime | LR | Selected weight | +0.19 (-3.56 to +1.61); 2/1/10; 13/15 analyses | +0.41 (-0.19 to +1.36); 2/0/3 |
| Stanford | cefepime | LR | Class-adjusted | +0.95 (-4.55 to +2.59); 2/1/12 | +0.49 (+0.34 to +2.13); 1/0/4 |
| Stanford | cefepime | LR | Prior-corrected | +0.00 (-5.52 to +1.62); 0/1/14 | +0.37 (+0.09 to +1.95); 1/0/4 |
| Stanford | cefepime | LR | Equal weight | +1.22 (-2.77 to +3.32); 3/0/12 | +1.25 (+0.18 to +1.39); 2/0/3 |
| Stanford | cefepime | HGB | Selected weight | +1.07 (-0.83 to +2.60); 6/1/6; 13/15 analyses | +0.15 (-0.36 to +2.10); 2/0/3 |
| Stanford | cefepime | HGB | Class-adjusted | +0.42 (-1.03 to +2.14); 3/0/12 | +0.26 (-0.67 to +1.91); 1/0/4 |
| Stanford | cefepime | HGB | Prior-corrected | -0.81 (-2.12 to +1.67); 0/2/13 | +0.20 (-0.61 to +1.98); 0/0/5 |
| Stanford | cefepime | HGB | Equal weight | +1.62 (+0.09 to +3.81); 8/0/7 | +0.78 (+0.41 to +2.34); 2/0/3 |
| Stanford | cefoxitin | LR | Selected weight | +0.41 (-0.44 to +10.05); 5/0/10 | +0.18 (+0.01 to +0.25); 1/0/4 |
| Stanford | cefoxitin | LR | Class-adjusted | +6.97 (+2.39 to +16.07); 15/0/0 | +1.14 (+0.97 to +2.29); 3/0/2 |
| Stanford | cefoxitin | LR | Prior-corrected | +6.36 (+1.80 to +15.71); 14/0/1 | +1.07 (+0.91 to +2.16); 3/0/2 |
| Stanford | cefoxitin | LR | Equal weight | +8.86 (+5.07 to +12.14); 15/0/0 | +2.00 (+1.55 to +2.16); 5/0/0 |
| Stanford | cefoxitin | HGB | Selected weight | +1.12 (-0.19 to +2.63); 7/0/8 | +0.18 (-0.16 to +0.56); 0/0/5 |
| Stanford | cefoxitin | HGB | Class-adjusted | +3.90 (+0.19 to +9.55); 11/0/4 | +0.80 (+0.33 to +1.10); 2/0/3 |
| Stanford | cefoxitin | HGB | Prior-corrected | +4.76 (+0.70 to +11.37); 11/0/4 | +0.70 (+0.35 to +1.18); 2/0/3 |
| Stanford | cefoxitin | HGB | Equal weight | +9.21 (+4.34 to +13.53); 15/0/0 | +1.68 (+1.26 to +2.43); 5/0/0 |
| Stanford | ceftazidime | LR | Selected weight | +1.39 (-0.38 to +3.85); 10/0/5 | -0.00 (-0.08 to +0.32); 1/0/4 |
| Stanford | ceftazidime | LR | Class-adjusted | +3.19 (+0.21 to +5.33); 14/0/1 | +0.49 (+0.08 to +0.71); 3/0/2 |
| Stanford | ceftazidime | LR | Prior-corrected | +3.15 (-0.04 to +5.00); 14/0/1 | +0.46 (+0.04 to +0.72); 2/0/3 |
| Stanford | ceftazidime | LR | Equal weight | +3.66 (+1.76 to +7.62); 14/0/1 | +0.50 (+0.07 to +0.89); 4/0/1 |
| Stanford | ceftazidime | HGB | Selected weight | +1.30 (-0.32 to +4.32); 9/0/6 | +0.26 (-0.10 to +0.51); 2/0/3 |
| Stanford | ceftazidime | HGB | Class-adjusted | +1.80 (+0.84 to +3.20); 9/0/6 | +0.41 (+0.11 to +1.00); 2/0/3 |
| Stanford | ceftazidime | HGB | Prior-corrected | +2.61 (+1.27 to +3.59); 13/0/2 | +0.50 (+0.10 to +1.05); 2/0/3 |
| Stanford | ceftazidime | HGB | Equal weight | +3.32 (+0.19 to +7.27); 13/0/2 | +0.87 (+0.18 to +1.10); 4/0/1 |
| UTSW | cefepime | LR | Selected weight | +0.11 (-0.74 to +0.80); 4/0/10; 14/15 analyses | +0.02 (+0.00 to +0.06); 0/0/5 |
| UTSW | cefepime | LR | Class-adjusted | +0.47 (-0.05 to +0.98); 11/0/4 | +0.07 (+0.01 to +0.10); 1/0/4 |
| UTSW | cefepime | LR | Prior-corrected | +0.26 (-0.09 to +0.76); 6/0/9 | +0.06 (+0.01 to +0.10); 1/0/4 |
| UTSW | cefepime | LR | Equal weight | +2.53 (+0.55 to +4.02); 14/0/1 | +0.71 (+0.31 to +1.33); 3/0/2 |
| UTSW | cefepime | HGB | Selected weight | +0.13 (-0.98 to +0.76); 5/0/9; 14/15 analyses | +0.01 (-0.01 to +0.06); 0/0/5 |
| UTSW | cefepime | HGB | Class-adjusted | +0.40 (-0.08 to +0.95); 8/0/7 | +0.06 (+0.01 to +0.10); 1/0/4 |
| UTSW | cefepime | HGB | Prior-corrected | +0.39 (-0.11 to +0.93); 8/0/7 | +0.06 (+0.01 to +0.10); 1/0/4 |
| UTSW | cefepime | HGB | Equal weight | +2.49 (+0.28 to +4.01); 14/0/1 | +0.71 (+0.31 to +1.33); 3/0/2 |
| UTSW | ceftazidime | LR | Selected weight | +1.10 (-5.10 to +114.76); 4/1/10 | +0.49 (-1.33 to +0.64); 0/0/5 |
| UTSW | ceftazidime | LR | Class-adjusted | +5.18 (-4.97 to +16.97); 7/1/7 | +1.22 (-1.36 to +2.05); 1/0/4 |
| UTSW | ceftazidime | LR | Prior-corrected | +1.84 (-6.77 to +11.15); 3/2/10 | +1.07 (-1.41 to +1.93); 1/0/4 |
| UTSW | ceftazidime | LR | Equal weight | +126.51 (+119.73 to +173.35); 15/0/0 | +108.73 (+101.38 to +149.18); 5/0/0 |
| UTSW | ceftazidime | HGB | Selected weight | -1.34 (-8.66 to +85.91); 3/1/11 | +0.47 (-1.35 to +0.67); 0/0/5 |
| UTSW | ceftazidime | HGB | Class-adjusted | +9.31 (-4.56 to +18.28); 9/0/6 | +1.16 (-1.39 to +2.06); 1/0/4 |
| UTSW | ceftazidime | HGB | Prior-corrected | +8.64 (-6.42 to +15.01); 9/0/6 | +1.16 (-1.39 to +2.06); 1/0/4 |
| UTSW | ceftazidime | HGB | Equal weight | +131.16 (+120.56 to +174.41); 15/0/0 | +108.65 (+101.30 to +149.12); 5/0/0 |

The selected weight did not consistently improve on the all-budget baseline. Its median difference was positive in 38 of the 40 drug, model and budget summaries; the two exceptions were UT Southwestern ceftazidime at 10% (gradient boosting, −1.34 × 10^−3^) and Stanford ceftazidime at 100% (logistic regression, −0.003 × 10^−3^). At the 10% budget the medians (× 10^−3^, logistic regression / gradient boosting) were +0.19 / +1.07 for Stanford cefepime, +0.41 / +1.12 for cefoxitin and +1.39 / +1.30 for ceftazidime, and +0.11 / +0.13 for UT Southwestern cefepime and +1.10 / −1.34 for ceftazidime. Most individual intervals included zero. Across all budgets, the intervals lay above zero in 100 of 296 Stanford analyses and 31 of 198 UT Southwestern analyses, and below zero in 3 and 5. The direction varied between splits for UT Southwestern ceftazidime at 10%: for gradient boosting, 3 of 5 split means were below zero. Results there were also unstable, with single analyses reaching +0.115 for logistic regression; the tuning sets in these analyses contained about 31 cultures. Class adjustment and prior correction were also mostly worse than the all-budget baseline (Table 4).

The tuning set often selected a weight of zero, which reproduces the reported-only model: in 146 of 296 Stanford analyses and 153 of 198 UT Southwestern analyses (Supplementary Table S3). For the selected weight, the comparison with the all-budget baseline then largely reflects the reported labels held out for tuning and calibration. The all-budget baseline itself had a lower median Brier score than the reported-only model in every Stanford summary. At UT Southwestern, differences between these two were small and mixed in sign.

### 3.5 Discrimination and recalibration

Rule-derived labels changed AUROC little relative to the reported-only and all-budget models (Figure 4). At Stanford, the median AUROC of logistic regression at the 100% budget was 0.70 for cefepime, 0.67 for cefoxitin and 0.71 for ceftazidime. At UT Southwestern, the age-only models were close to chance: median AUROC 0.49 to 0.54 for cefepime and 0.49 to 0.53 for ceftazidime across budgets and methods. The UT Southwestern comparisons are therefore informative mainly about calibration and the effect of class balance, not about ranking.

**Figure 4.**
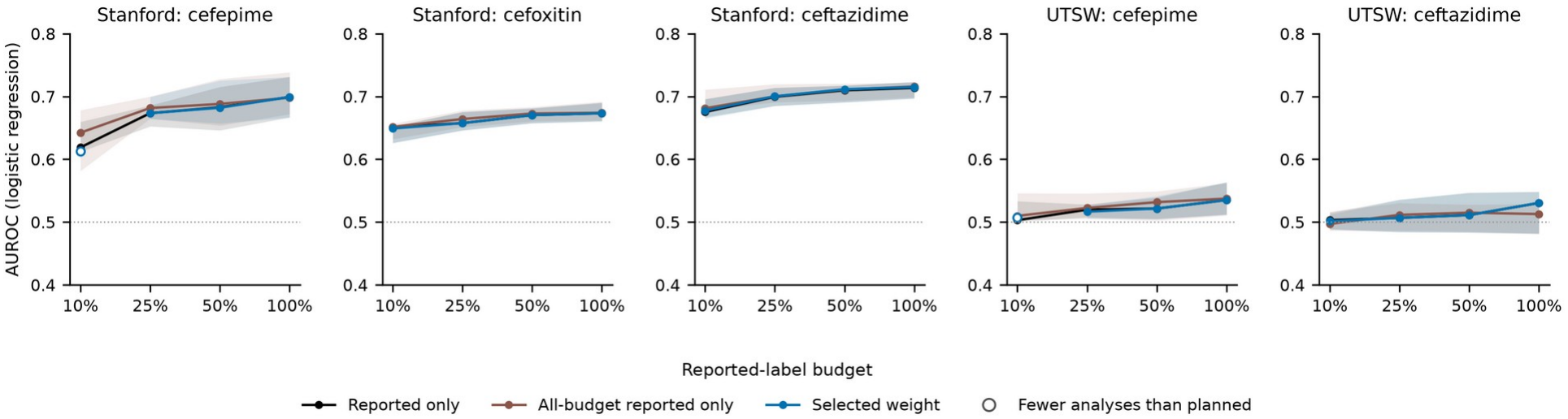
AUROC by reported-label budget (logistic regression). Line: median across outer splits of the split mean; band: range across outer splits; hollow points rest on fewer analyses than planned. Dotted line: AUROC 0.5.

Recalibration was often unavailable at small budgets because calibration sets had too few positives. For Stanford cefepime at 10%, the recalibrated reported-only model was available in 3 of 15 analyses and the recalibrated selected-weight comparison in 2 of 15. Where both recalibrated models were available, their differences were usually exactly zero, because the tuning set had selected α = 0. Remaining differences were small and of mixed sign (exported results).

Median training times per model at the 100% budget were below 0.4 s for every logistic regression method and below 0.35 s for gradient boosting methods without weight selection. The selected weight required several fits (median 1.8 s for gradient boosting at Stanford).

## 4. Discussion

At two sites, three antibiotics and two model classes, adding rule-derived labels did not consistently improve probability prediction over a reported-only model that used the entire reported-label budget for training. Equal-weight augmentation was clearly worse, and the tuning-selected weight was usually slightly worse or indistinguishable. These results do not show that rule-derived labels cannot help. They show that, in this design, any benefit was not large enough to offset the reported labels set aside for weight selection, and that unweighted addition of single-class labels damaged calibration.

The result resembles Zhu et al.’s observation that clean validation data are often better spent on training [4], but the settings differ: tabular features, a small number of rules, labels that only ever imply the majority class, and probability-based evaluation. Our study does not test whether their conclusions hold generally. It suggests that the comparison they recommend, against a model trained on all the clean labels, is also informative in this application. Choosing α on a tuning set is a coarse form of the validation-guided reweighting studied by Ren et al. [5]. That α = 0 was chosen in about half of the Stanford analyses and three-quarters of the UT Southwestern analyses indicates that the tuning sets often found no benefit from the rule-derived records.

The calibration findings agree with work on class-imbalance correction in risk prediction [7,8]. Adding many susceptible labels acts like oversampling the majority class, and the resulting underestimation of risk could largely be undone by restoring the reported class proportions, either through weights or through a prior correction [11]. These corrections recovered calibration-in-the-large but not the Brier score of the all-budget baseline. A likely contributor is that rule-derived cultures differ from reported cultures in more than class proportion, violating the label-shift assumption [10]. We did not test this directly.

Weak-label quality is uncertain at both sites. Stanford’s implied labels follow documented laboratory protocols [12], but we know of no independent validation of their accuracy. At UT Southwestern, the labels are our own application of Stanford rule text. Agreement with reported results was high on co-tested cultures (for example, 16 disagreements among 16,690 cefepime cultures), but those cultures were selected for testing and may not represent the cultures that received weak labels. Our check on Stanford data also showed that the rule text alone does not reproduce Stanford’s released labels. WRENCH highlighted that weak-label sources differ in ways that complicate comparison [2]. The same applies here: the two sites differ in how labels were produced as well as in patients, testing practice and features.

The two-site design should not be read as external validation. Following the distinction drawn in prediction-model research [14], we did not take a model fitted at Stanford and evaluate it at UT Southwestern. We repeated the design and refitted every model at each site. The primary feature sets differ (rich at Stanford, age-only at UT Southwestern), and the UT Southwestern models had almost no discrimination. The UT Southwestern results therefore test whether the label-budget findings recur under weaker features and differently built labels, not whether a particular model transports. We also could not verify whether the two datasets share patients.

### Strengths and limitations

Strengths include a design in which every method uses the same reported patients within an allocation; an explicit all-budget baseline; separate tuning and calibration patients; paired cluster bootstrap intervals; reporting of availability, including analyses that could not be run; and checks that a selected weight of zero reproduces the reported-only model exactly.

The limitations are substantial. The analysis is exploratory, and the Stanford data had been examined in earlier analyses with the same outer splits, so there is no untouched confirmatory test set. Outer splits overlap, and we made many comparisons without adjustment. Bootstrap intervals cover only test-patient sampling. Reported results may be affected by selective testing and are not assumed to be error-free; the UT Southwestern ceftazidime prevalence of 38% suggests strong selection. The timing of resistance-history records is not verified, so leakage of information unavailable at prediction time cannot be excluded at Stanford. Dates are jittered, so no temporal validation was attempted. The budgets were simulated by subsampling fully labelled data, and real low-label settings may differ in which cultures are labelled. Hyperparameters were fixed. The rules imply only susceptibility. The study does not assess clinical utility, and nothing here supports clinical use of these models or rules. A preliminary feature-matched comparison between sites was not part of the completed analyses and is not reported.

### Future work

Useful next steps are a prespecified analysis on data not used during method development; feature sets that are matched across sites where meanings can be established; weak-label sources that can imply both classes; and an independent assessment of rule validity at each laboratory. Any new method for combining weak and reported labels should be compared with an all-budget reported-only baseline and evaluated on calibration as well as discrimination.

## 5. Conclusion

In exploratory label-budget experiments at two sites, rule-derived susceptibility labels did not consistently improve probability prediction compared with using the whole reported-label budget for training, and unweighted addition of these single-class labels seriously miscalibrated predictions. Evaluations of weak supervision should count the reported labels used for tuning and calibration, include an all-budget reported-only baseline, and report calibration alongside discrimination.

## Data Availability

This study used two publicly available, de-identified datasets: the Stanford Antibiotic Resistance Microbiology Dataset (ARMD), available at https://doi.org/10.5061/dryad.jq2bvq8kp, and the Antimicrobial Resistance Microbiological Dataset from UT Southwestern (ARMD-UTSW), available at https://doi.org/10.5061/dryad.0rxwdbsd5. Aggregate results generated in this study are available from the corresponding author upon reasonable request. No patient-level data, identifiers, or predictions generated by this study are shared.

https://doi.org/10.5061/dryad.jq2bvq8kp

## Declarations

### Data availability

The source datasets were released by their providers and are available from them: Stanford ARMD (Dryad, doi:10.5061/dryad.jq2bvq8kp) [12] and ARMD-UTSW (Dryad, doi:10.5061/dryad.0rxwdbsd5) [13]. The aggregate results generated in this study are available from the corresponding author on request. No patient-level data, identifiers or predictions are shared.

### Code availability

The analysis notebooks (ARMD_label_budget_study_v4.ipynb for Stanford and ARMD_UTSW_final_study.ipynb for UT Southwestern), which record configuration, seeds, package versions and input-file hashes, are available from the corresponding author on request.

### Ethics

This study is a secondary analysis of publicly released, de-identified datasets; no additional ethical approval was required. The ARMD-UTSW documentation states that dataset creation was approved by the UT Southwestern IRB (#STU-2023-0583) and that patient consent was not required [13]. The Stanford dataset is described in its data descriptor [12].

### Author contributions

Talim Hossain Sadi: conceptualization, methodology, formal analysis, investigation, writing (original draft). Chowdhury Aseer Ruthbah: data curation, software, investigation, writing (original draft, supporting). All authors reviewed and approved the final manuscript.

---

**Supplementary Table S1.** Reported-label budget and its allocation: patients / cultures / non-susceptible cultures, median across outer splits and budget samples.

| Site | Drug | Budget | Budget total | Training | Tuning | Calibration |
|---|---|---|---|---|---|---|
| Stanford | cefepime | 10% | 982 / 1309 / 56 | 735 / 973 / 41 | 125 / 164 / 6 | 122 / 156 / 7 |
| Stanford | cefepime | 25% | 2455 / 3222 / 141 | 1850 / 2411 / 106 | 314 / 420 / 17 | 304 / 399 / 16 |
| Stanford | cefepime | 50% | 4909 / 6496 / 293 | 3676 / 4867 / 211 | 616 / 823 / 39 | 615 / 804 / 36 |
| Stanford | cefepime | 100% | 9818 / 13033 / 566 | 7390 / 9805 / 428 | 1234 / 1639 / 74 | 1232 / 1610 / 76 |
| Stanford | cefoxitin | 10% | 1592 / 2411 / 298 | 1191 / 1823 / 223 | 201 / 294 / 35 | 202 / 304 / 37 |
| Stanford | cefoxitin | 25% | 3978 / 5978 / 728 | 2988 / 4509 / 550 | 503 / 738 / 87 | 483 / 747 / 87 |
| Stanford | cefoxitin | 50% | 7956 / 12015 / 1427 | 5975 / 9034 / 1062 | 987 / 1503 / 187 | 995 / 1508 / 164 |
| Stanford | cefoxitin | 100% | 15911 / 23983 / 2827 | 11950 / 18058 / 2125 | 1984 / 3028 / 353 | 1993 / 2997 / 340 |
| Stanford | ceftazidime | 10% | 2003 / 2972 / 231 | 1507 / 2233 / 160 | 250 / 361 / 31 | 246 / 365 / 29 |
| Stanford | ceftazidime | 25% | 5007 / 7424 / 587 | 3768 / 5605 / 424 | 624 / 926 / 80 | 608 / 924 / 73 |
| Stanford | ceftazidime | 50% | 10013 / 14892 / 1181 | 7522 / 11237 / 878 | 1236 / 1852 / 148 | 1251 / 1866 / 149 |
| Stanford | ceftazidime | 100% | 20025 / 29843 / 2340 | 14993 / 22375 / 1728 | 2465 / 3743 / 312 | 2500 / 3692 / 285 |
| UTSW | cefepime | 10% | 1129 / 1584 / 97 | 850 / 1194 / 71 | 134 / 184 / 11 | 145 / 204 / 13 |
| UTSW | cefepime | 25% | 2821 / 3968 / 240 | 2120 / 2963 / 178 | 351 / 493 / 32 | 348 / 504 / 28 |
| UTSW | cefepime | 50% | 5641 / 7904 / 482 | 4251 / 5945 / 352 | 696 / 980 / 63 | 699 / 985 / 56 |
| UTSW | cefepime | 100% | 11282 / 15844 / 963 | 8498 / 11897 / 724 | 1404 / 1973 / 125 | 1405 / 1979 / 114 |
| UTSW | ceftazidime | 10% | 221 / 274 / 104 | 168 / 202 / 73 | 25 / 31 / 11 | 26 / 31 / 13 |
| UTSW | ceftazidime | 25% | 551 / 696 / 259 | 416 / 517 / 186 | 67 / 85 / 33 | 65 / 79 / 30 |
| UTSW | ceftazidime | 50% | 1101 / 1389 / 517 | 834 / 1043 / 385 | 136 / 175 / 68 | 133 / 160 / 62 |
| UTSW | ceftazidime | 100% | 2202 / 2767 / 1051 | 1669 / 2090 / 776 | 275 / 358 / 135 | 269 / 330 / 128 |

**Supplementary Table S2.**
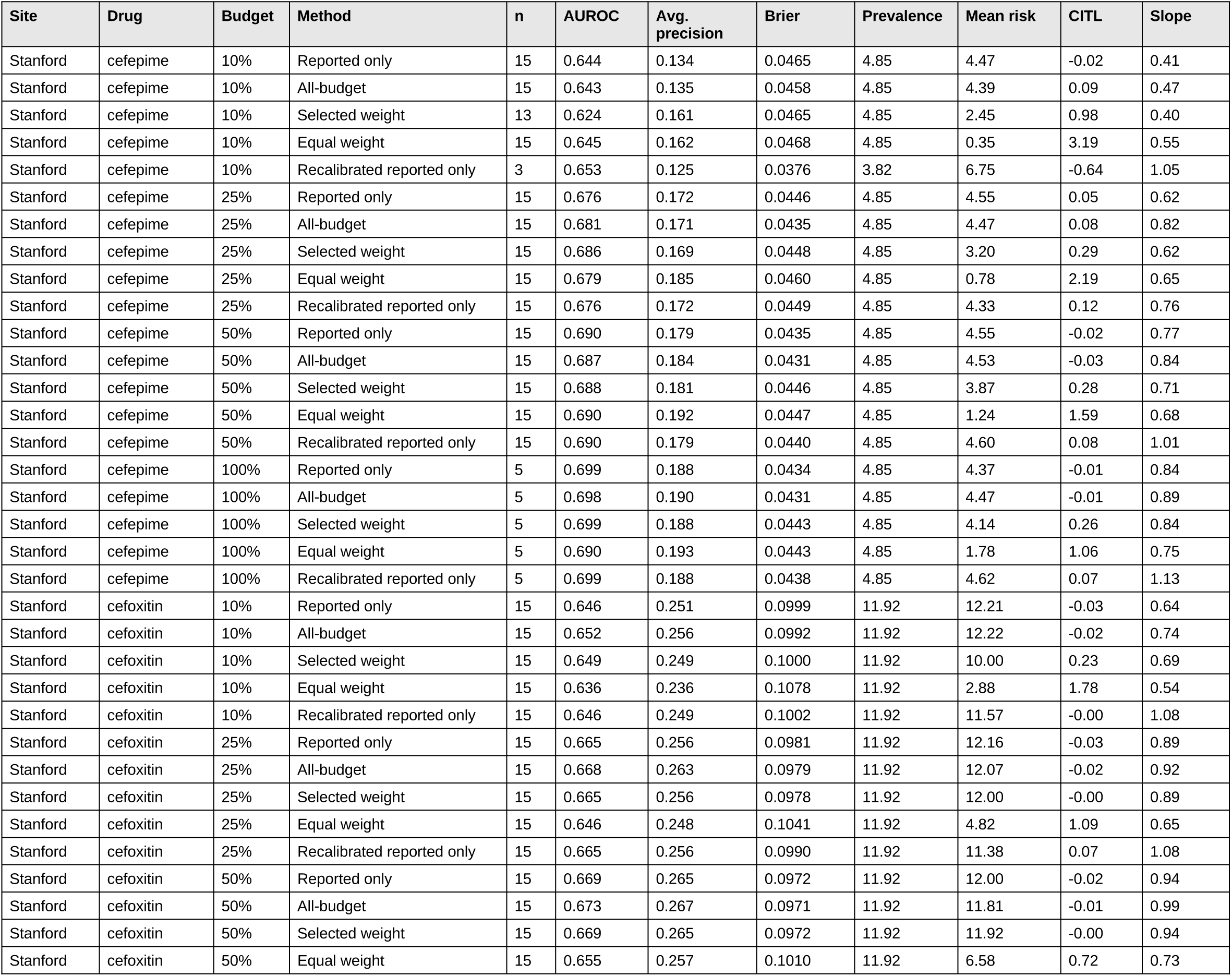
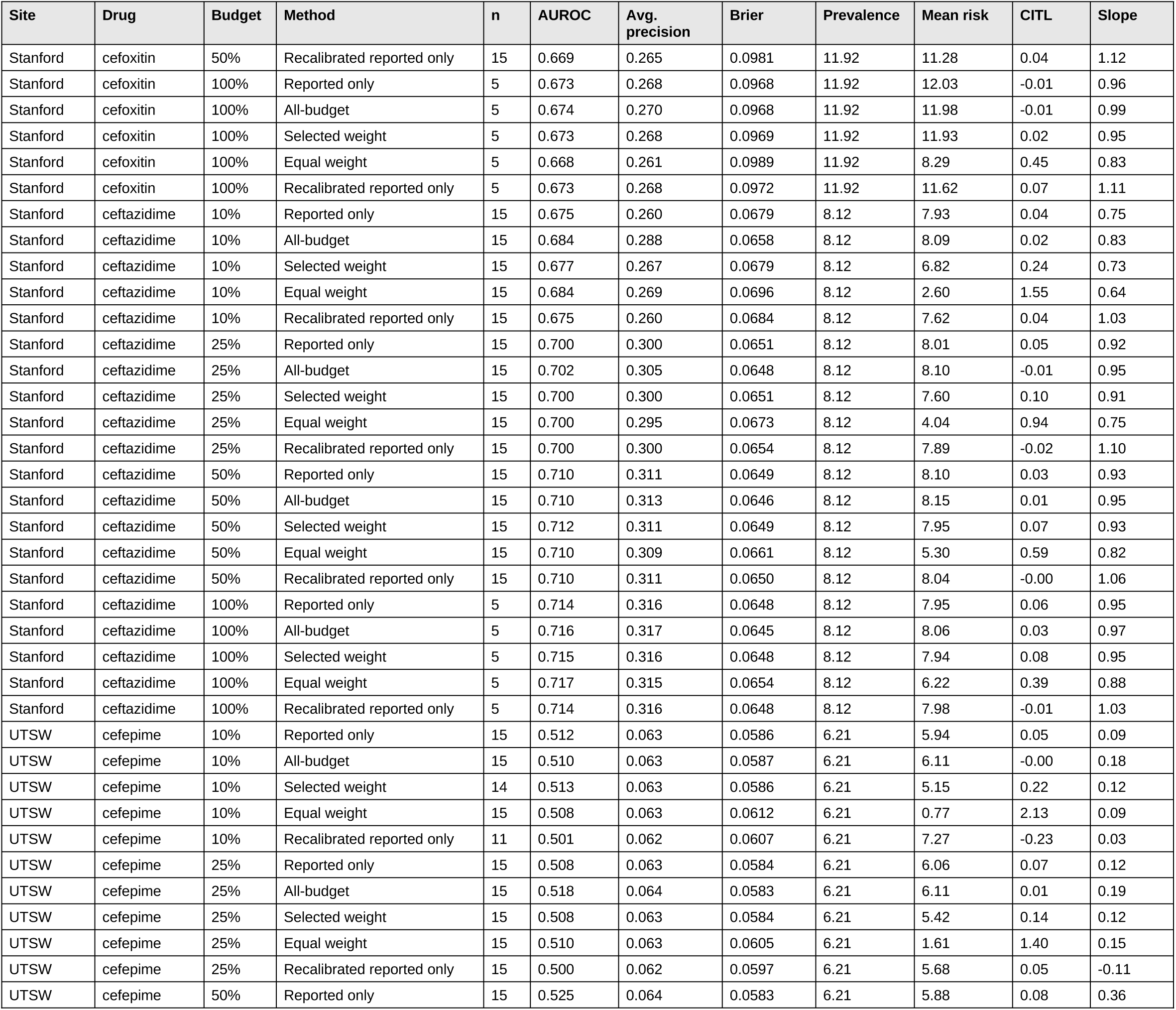
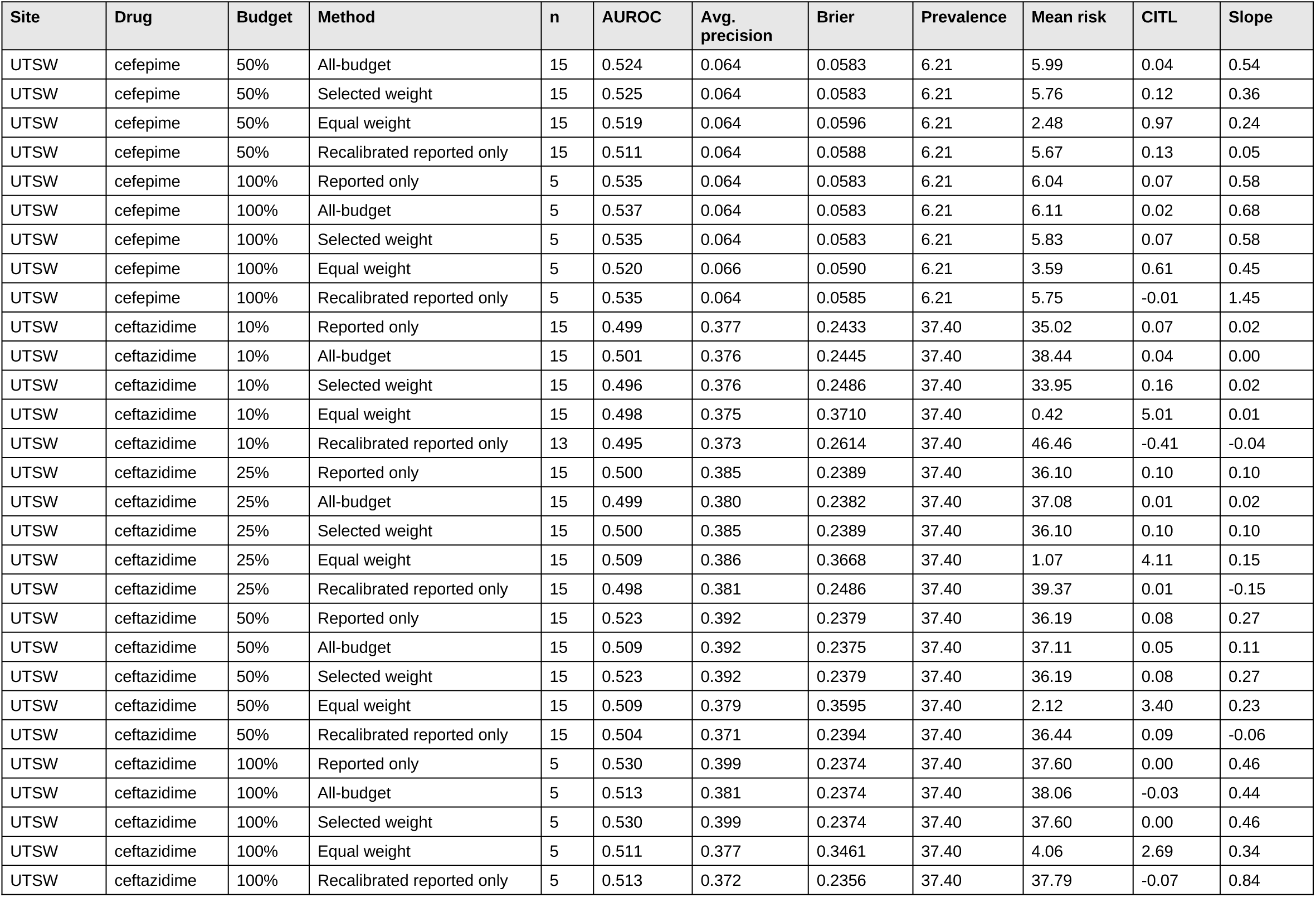
Test metrics for logistic regression: medians across available analyses (n). Prevalence and mean risk in %. CITL, calibration-in-the-large. Gradient-boosting results are in the exported files.

| Site | Drug | Budget | Method | n | AUROC | Avg. precision | Brier | Prevalence | Mean risk | CITL | Slope |
|---|---|---|---|---|---|---|---|---|---|---|---|
| Stanford | cefepime | 10% | Reported only | 15 | 0.644 | 0.134 | 0.0465 | 4.85 | 4.47 | -0.02 | 0.41 |
| Stanford | cefepime | 10% | All-budget | 15 | 0.643 | 0.135 | 0.0458 | 4.85 | 4.39 | 0.09 | 0.47 |
| Stanford | cefepime | 10% | Selected weight | 13 | 0.624 | 0.161 | 0.0465 | 4.85 | 2.45 | 0.98 | 0.40 |
| Stanford | cefepime | 10% | Equal weight | 15 | 0.645 | 0.162 | 0.0468 | 4.85 | 0.35 | 3.19 | 0.55 |
| Stanford | cefepime | 10% | Recalibrated reported only | 3 | 0.653 | 0.125 | 0.0376 | 3.82 | 6.75 | -0.64 | 1.05 |
| Stanford | cefepime | 25% | Reported only | 15 | 0.676 | 0.172 | 0.0446 | 4.85 | 4.55 | 0.05 | 0.62 |
| Stanford | cefepime | 25% | All-budget | 15 | 0.681 | 0.171 | 0.0435 | 4.85 | 4.47 | 0.08 | 0.82 |
| Stanford | cefepime | 25% | Selected weight | 15 | 0.686 | 0.169 | 0.0448 | 4.85 | 3.20 | 0.29 | 0.62 |
| Stanford | cefepime | 25% | Equal weight | 15 | 0.679 | 0.185 | 0.0460 | 4.85 | 0.78 | 2.19 | 0.65 |
| Stanford | cefepime | 25% | Recalibrated reported only | 15 | 0.676 | 0.172 | 0.0449 | 4.85 | 4.33 | 0.12 | 0.76 |
| Stanford | cefepime | 50% | Reported only | 15 | 0.690 | 0.179 | 0.0435 | 4.85 | 4.55 | -0.02 | 0.77 |
| Stanford | cefepime | 50% | All-budget | 15 | 0.687 | 0.184 | 0.0431 | 4.85 | 4.53 | -0.03 | 0.84 |
| Stanford | cefepime | 50% | Selected weight | 15 | 0.688 | 0.181 | 0.0446 | 4.85 | 3.87 | 0.28 | 0.71 |
| Stanford | cefepime | 50% | Equal weight | 15 | 0.690 | 0.192 | 0.0447 | 4.85 | 1.24 | 1.59 | 0.68 |
| Stanford | cefepime | 50% | Recalibrated reported only | 15 | 0.690 | 0.179 | 0.0440 | 4.85 | 4.60 | 0.08 | 1.01 |
| Stanford | cefepime | 100% | Reported only | 5 | 0.699 | 0.188 | 0.0434 | 4.85 | 4.37 | -0.01 | 0.84 |
| Stanford | cefepime | 100% | All-budget | 5 | 0.698 | 0.190 | 0.0431 | 4.85 | 4.47 | -0.01 | 0.89 |
| Stanford | cefepime | 100% | Selected weight | 5 | 0.699 | 0.188 | 0.0443 | 4.85 | 4.14 | 0.26 | 0.84 |
| Stanford | cefepime | 100% | Equal weight | 5 | 0.690 | 0.193 | 0.0443 | 4.85 | 1.78 | 1.06 | 0.75 |
| Stanford | cefepime | 100% | Recalibrated reported only | 5 | 0.699 | 0.188 | 0.0438 | 4.85 | 4.62 | 0.07 | 1.13 |
| Stanford | cefoxitin | 10% | Reported only | 15 | 0.646 | 0.251 | 0.0999 | 11.92 | 12.21 | -0.03 | 0.64 |
| Stanford | cefoxitin | 10% | All-budget | 15 | 0.652 | 0.256 | 0.0992 | 11.92 | 12.22 | -0.02 | 0.74 |
| Stanford | cefoxitin | 10% | Selected weight | 15 | 0.649 | 0.249 | 0.1000 | 11.92 | 10.00 | 0.23 | 0.69 |
| Stanford | cefoxitin | 10% | Equal weight | 15 | 0.636 | 0.236 | 0.1078 | 11.92 | 2.88 | 1.78 | 0.54 |
| Stanford | cefoxitin | 10% | Recalibrated reported only | 15 | 0.646 | 0.249 | 0.1002 | 11.92 | 11.57 | -0.00 | 1.08 |
| Stanford | cefoxitin | 25% | Reported only | 15 | 0.665 | 0.256 | 0.0981 | 11.92 | 12.16 | -0.03 | 0.89 |
| Stanford | cefoxitin | 25% | All-budget | 15 | 0.668 | 0.263 | 0.0979 | 11.92 | 12.07 | -0.02 | 0.92 |
| Stanford | cefoxitin | 25% | Selected weight | 15 | 0.665 | 0.256 | 0.0978 | 11.92 | 12.00 | -0.00 | 0.89 |
| Stanford | cefoxitin | 25% | Equal weight | 15 | 0.646 | 0.248 | 0.1041 | 11.92 | 4.82 | 1.09 | 0.65 |
| Stanford | cefoxitin | 25% | Recalibrated reported only | 15 | 0.665 | 0.256 | 0.0990 | 11.92 | 11.38 | 0.07 | 1.08 |
| Stanford | cefoxitin | 50% | Reported only | 15 | 0.669 | 0.265 | 0.0972 | 11.92 | 12.00 | -0.02 | 0.94 |
| Stanford | cefoxitin | 50% | All-budget | 15 | 0.673 | 0.267 | 0.0971 | 11.92 | 11.81 | -0.01 | 0.99 |
| Stanford | cefoxitin | 50% | Selected weight | 15 | 0.669 | 0.265 | 0.0972 | 11.92 | 11.92 | -0.00 | 0.94 |
| Stanford | cefoxitin | 50% | Equal weight | 15 | 0.655 | 0.257 | 0.1010 | 11.92 | 6.58 | 0.72 | 0.73 |
| Site | Drug | Budget | Method | n | AUROC | Avg. precision | Brier | Prevalence | Mean risk | CITL | Slope |
| Stanford | cefoxitin | 50% | Recalibrated reported only | 15 | 0.669 | 0.265 | 0.0981 | 11.92 | 11.28 | 0.04 | 1.12 |
| Stanford | cefoxitin | 100% | Reported only | 5 | 0.673 | 0.268 | 0.0968 | 11.92 | 12.03 | -0.01 | 0.96 |
| Stanford | cefoxitin | 100% | All-budget | 5 | 0.674 | 0.270 | 0.0968 | 11.92 | 11.98 | -0.01 | 0.99 |
| Stanford | cefoxitin | 100% | Selected weight | 5 | 0.673 | 0.268 | 0.0969 | 11.92 | 11.93 | 0.02 | 0.95 |
| Stanford | cefoxitin | 100% | Equal weight | 5 | 0.668 | 0.261 | 0.0989 | 11.92 | 8.29 | 0.45 | 0.83 |
| Stanford | cefoxitin | 100% | Recalibrated reported only | 5 | 0.673 | 0.268 | 0.0972 | 11.92 | 11.62 | 0.07 | 1.11 |
| Stanford | ceftazidime | 10% | Reported only | 15 | 0.675 | 0.260 | 0.0679 | 8.12 | 7.93 | 0.04 | 0.75 |
| Stanford | ceftazidime | 10% | All-budget | 15 | 0.684 | 0.288 | 0.0658 | 8.12 | 8.09 | 0.02 | 0.83 |
| Stanford | ceftazidime | 10% | Selected weight | 15 | 0.677 | 0.267 | 0.0679 | 8.12 | 6.82 | 0.24 | 0.73 |
| Stanford | ceftazidime | 10% | Equal weight | 15 | 0.684 | 0.269 | 0.0696 | 8.12 | 2.60 | 1.55 | 0.64 |
| Stanford | ceftazidime | 10% | Recalibrated reported only | 15 | 0.675 | 0.260 | 0.0684 | 8.12 | 7.62 | 0.04 | 1.03 |
| Stanford | ceftazidime | 25% | Reported only | 15 | 0.700 | 0.300 | 0.0651 | 8.12 | 8.01 | 0.05 | 0.92 |
| Stanford | ceftazidime | 25% | All-budget | 15 | 0.702 | 0.305 | 0.0648 | 8.12 | 8.10 | -0.01 | 0.95 |
| Stanford | ceftazidime | 25% | Selected weight | 15 | 0.700 | 0.300 | 0.0651 | 8.12 | 7.60 | 0.10 | 0.91 |
| Stanford | ceftazidime | 25% | Equal weight | 15 | 0.700 | 0.295 | 0.0673 | 8.12 | 4.04 | 0.94 | 0.75 |
| Stanford | ceftazidime | 25% | Recalibrated reported only | 15 | 0.700 | 0.300 | 0.0654 | 8.12 | 7.89 | -0.02 | 1.10 |
| Stanford | ceftazidime | 50% | Reported only | 15 | 0.710 | 0.311 | 0.0649 | 8.12 | 8.10 | 0.03 | 0.93 |
| Stanford | ceftazidime | 50% | All-budget | 15 | 0.710 | 0.313 | 0.0646 | 8.12 | 8.15 | 0.01 | 0.95 |
| Stanford | ceftazidime | 50% | Selected weight | 15 | 0.712 | 0.311 | 0.0649 | 8.12 | 7.95 | 0.07 | 0.93 |
| Stanford | ceftazidime | 50% | Equal weight | 15 | 0.710 | 0.309 | 0.0661 | 8.12 | 5.30 | 0.59 | 0.82 |
| Stanford | ceftazidime | 50% | Recalibrated reported only | 15 | 0.710 | 0.311 | 0.0650 | 8.12 | 8.04 | -0.00 | 1.06 |
| Stanford | ceftazidime | 100% | Reported only | 5 | 0.714 | 0.316 | 0.0648 | 8.12 | 7.95 | 0.06 | 0.95 |
| Stanford | ceftazidime | 100% | All-budget | 5 | 0.716 | 0.317 | 0.0645 | 8.12 | 8.06 | 0.03 | 0.97 |
| Stanford | ceftazidime | 100% | Selected weight | 5 | 0.715 | 0.316 | 0.0648 | 8.12 | 7.94 | 0.08 | 0.95 |
| Stanford | ceftazidime | 100% | Equal weight | 5 | 0.717 | 0.315 | 0.0654 | 8.12 | 6.22 | 0.39 | 0.88 |
| Stanford | ceftazidime | 100% | Recalibrated reported only | 5 | 0.714 | 0.316 | 0.0648 | 8.12 | 7.98 | -0.01 | 1.03 |
| UTSW | cefepime | 10% | Reported only | 15 | 0.512 | 0.063 | 0.0586 | 6.21 | 5.94 | 0.05 | 0.09 |
| UTSW | cefepime | 10% | All-budget | 15 | 0.510 | 0.063 | 0.0587 | 6.21 | 6.11 | -0.00 | 0.18 |
| UTSW | cefepime | 10% | Selected weight | 14 | 0.513 | 0.063 | 0.0586 | 6.21 | 5.15 | 0.22 | 0.12 |
| UTSW | cefepime | 10% | Equal weight | 15 | 0.508 | 0.063 | 0.0612 | 6.21 | 0.77 | 2.13 | 0.09 |
| UTSW | cefepime | 10% | Recalibrated reported only | 11 | 0.501 | 0.062 | 0.0607 | 6.21 | 7.27 | -0.23 | 0.03 |
| UTSW | cefepime | 25% | Reported only | 15 | 0.508 | 0.063 | 0.0584 | 6.21 | 6.06 | 0.07 | 0.12 |
| UTSW | cefepime | 25% | All-budget | 15 | 0.518 | 0.064 | 0.0583 | 6.21 | 6.11 | 0.01 | 0.19 |
| UTSW | cefepime | 25% | Selected weight | 15 | 0.508 | 0.063 | 0.0584 | 6.21 | 5.42 | 0.14 | 0.12 |
| UTSW | cefepime | 25% | Equal weight | 15 | 0.510 | 0.063 | 0.0605 | 6.21 | 1.61 | 1.40 | 0.15 |
| UTSW | cefepime | 25% | Recalibrated reported only | 15 | 0.500 | 0.062 | 0.0597 | 6.21 | 5.68 | 0.05 | -0.11 |
| UTSW | cefepime | 50% | Reported only | 15 | 0.525 | 0.064 | 0.0583 | 6.21 | 5.88 | 0.08 | 0.36 |
| Site | Drug | Budget | Method | n | AUROC | Avg. precision | Brier | Prevalence | Mean risk | CITL | Slope |
| UTSW | cefepime | 50% | All-budget | 15 | 0.524 | 0.064 | 0.0583 | 6.21 | 5.99 | 0.04 | 0.54 |
| UTSW | cefepime | 50% | Selected weight | 15 | 0.525 | 0.064 | 0.0583 | 6.21 | 5.76 | 0.12 | 0.36 |
| UTSW | cefepime | 50% | Equal weight | 15 | 0.519 | 0.064 | 0.0596 | 6.21 | 2.48 | 0.97 | 0.24 |
| UTSW | cefepime | 50% | Recalibrated reported only | 15 | 0.511 | 0.064 | 0.0588 | 6.21 | 5.67 | 0.13 | 0.05 |
| UTSW | cefepime | 100% | Reported only | 5 | 0.535 | 0.064 | 0.0583 | 6.21 | 6.04 | 0.07 | 0.58 |
| UTSW | cefepime | 100% | All-budget | 5 | 0.537 | 0.064 | 0.0583 | 6.21 | 6.11 | 0.02 | 0.68 |
| UTSW | cefepime | 100% | Selected weight | 5 | 0.535 | 0.064 | 0.0583 | 6.21 | 5.83 | 0.07 | 0.58 |
| UTSW | cefepime | 100% | Equal weight | 5 | 0.520 | 0.066 | 0.0590 | 6.21 | 3.59 | 0.61 | 0.45 |
| UTSW | cefepime | 100% | Recalibrated reported only | 5 | 0.535 | 0.064 | 0.0585 | 6.21 | 5.75 | -0.01 | 1.45 |
| UTSW | ceftazidime | 10% | Reported only | 15 | 0.499 | 0.377 | 0.2433 | 37.40 | 35.02 | 0.07 | 0.02 |
| UTSW | ceftazidime | 10% | All-budget | 15 | 0.501 | 0.376 | 0.2445 | 37.40 | 38.44 | 0.04 | 0.00 |
| UTSW | ceftazidime | 10% | Selected weight | 15 | 0.496 | 0.376 | 0.2486 | 37.40 | 33.95 | 0.16 | 0.02 |
| UTSW | ceftazidime | 10% | Equal weight | 15 | 0.498 | 0.375 | 0.3710 | 37.40 | 0.42 | 5.01 | 0.01 |
| UTSW | ceftazidime | 10% | Recalibrated reported only | 13 | 0.495 | 0.373 | 0.2614 | 37.40 | 46.46 | -0.41 | -0.04 |
| UTSW | ceftazidime | 25% | Reported only | 15 | 0.500 | 0.385 | 0.2389 | 37.40 | 36.10 | 0.10 | 0.10 |
| UTSW | ceftazidime | 25% | All-budget | 15 | 0.499 | 0.380 | 0.2382 | 37.40 | 37.08 | 0.01 | 0.02 |
| UTSW | ceftazidime | 25% | Selected weight | 15 | 0.500 | 0.385 | 0.2389 | 37.40 | 36.10 | 0.10 | 0.10 |
| UTSW | ceftazidime | 25% | Equal weight | 15 | 0.509 | 0.386 | 0.3668 | 37.40 | 1.07 | 4.11 | 0.15 |
| UTSW | ceftazidime | 25% | Recalibrated reported only | 15 | 0.498 | 0.381 | 0.2486 | 37.40 | 39.37 | 0.01 | -0.15 |
| UTSW | ceftazidime | 50% | Reported only | 15 | 0.523 | 0.392 | 0.2379 | 37.40 | 36.19 | 0.08 | 0.27 |
| UTSW | ceftazidime | 50% | All-budget | 15 | 0.509 | 0.392 | 0.2375 | 37.40 | 37.11 | 0.05 | 0.11 |
| UTSW | ceftazidime | 50% | Selected weight | 15 | 0.523 | 0.392 | 0.2379 | 37.40 | 36.19 | 0.08 | 0.27 |
| UTSW | ceftazidime | 50% | Equal weight | 15 | 0.509 | 0.379 | 0.3595 | 37.40 | 2.12 | 3.40 | 0.23 |
| UTSW | ceftazidime | 50% | Recalibrated reported only | 15 | 0.504 | 0.371 | 0.2394 | 37.40 | 36.44 | 0.09 | -0.06 |
| UTSW | ceftazidime | 100% | Reported only | 5 | 0.530 | 0.399 | 0.2374 | 37.40 | 37.60 | 0.00 | 0.46 |
| UTSW | ceftazidime | 100% | All-budget | 5 | 0.513 | 0.381 | 0.2374 | 37.40 | 38.06 | -0.03 | 0.44 |
| UTSW | ceftazidime | 100% | Selected weight | 5 | 0.530 | 0.399 | 0.2374 | 37.40 | 37.60 | 0.00 | 0.46 |
| UTSW | ceftazidime | 100% | Equal weight | 5 | 0.511 | 0.377 | 0.3461 | 37.40 | 4.06 | 2.69 | 0.34 |
| UTSW | ceftazidime | 100% | Recalibrated reported only | 5 | 0.513 | 0.372 | 0.2356 | 37.40 | 37.79 | -0.07 | 0.84 |

**Supplementary Table S3.**
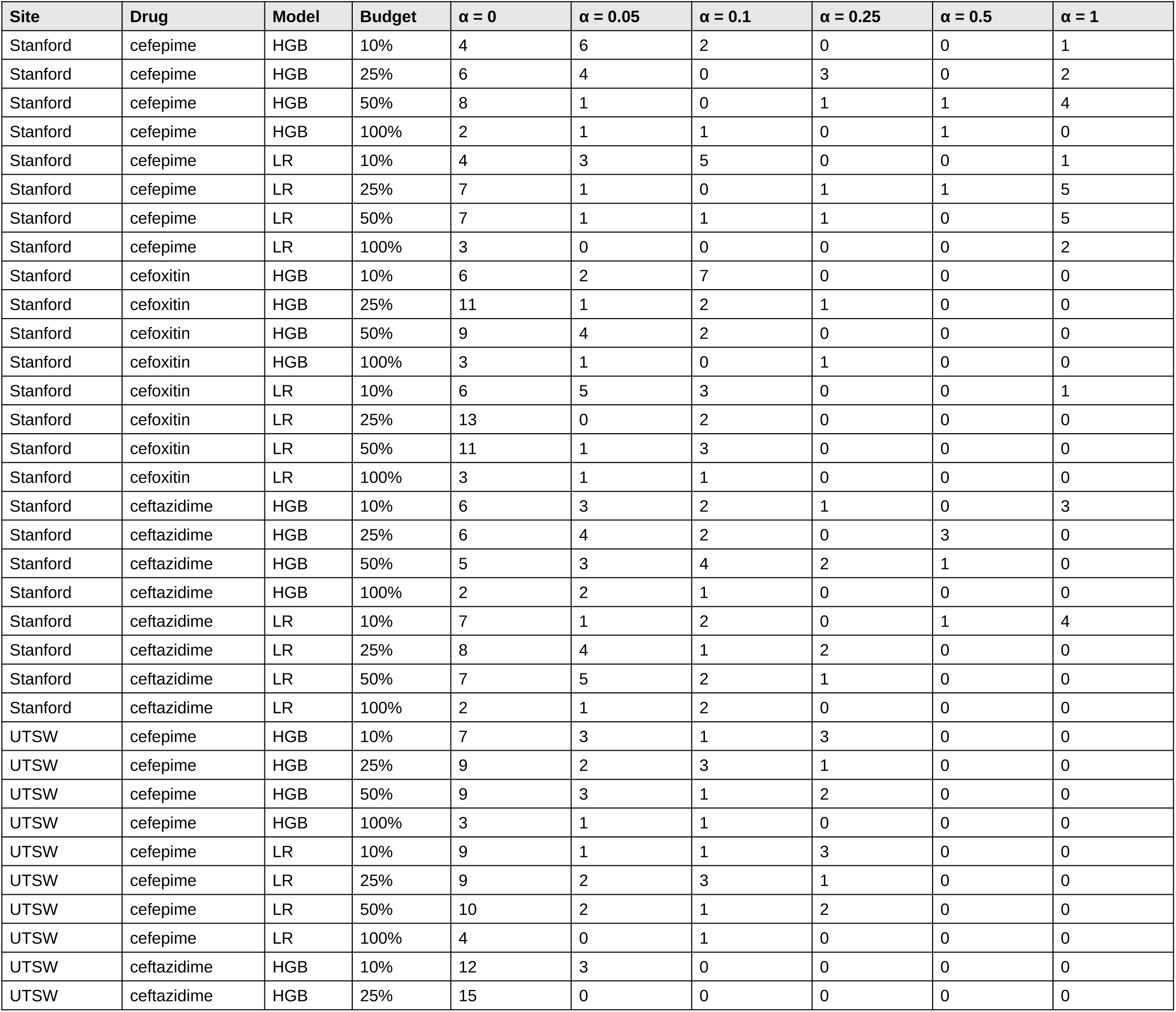
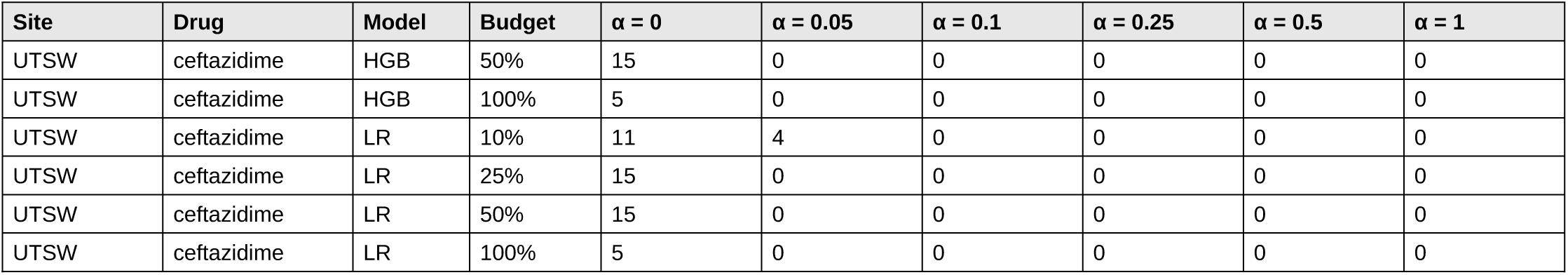
Inferred weight α selected on tuning cultures (number of analyses).

| **Site** | **Drug** | **Model** | **Budget** | **α = 0** | **α = 0.05** | **α = 0.1** | **α = 0.25** | **α = 0.5** | **α = 1** |
|---|---|---|---|---|---|---|---|---|---|
| Stanford | cefepime | HGB | 10% | 4 | 6 | 2 | 0 | 0 | 1 |
| Stanford | cefepime | HGB | 25% | 6 | 4 | 0 | 3 | 0 | 2 |
| Stanford | cefepime | HGB | 50% | 8 | 1 | 0 | 1 | 1 | 4 |
| Stanford | cefepime | HGB | 100% | 2 | 1 | 1 | 0 | 1 | 0 |
| Stanford | cefepime | LR | 10% | 4 | 3 | 5 | 0 | 0 | 1 |
| Stanford | cefepime | LR | 25% | 7 | 1 | 0 | 1 | 1 | 5 |
| Stanford | cefepime | LR | 50% | 7 | 1 | 1 | 1 | 0 | 5 |
| Stanford | cefepime | LR | 100% | 3 | 0 | 0 | 0 | 0 | 2 |
| Stanford | cefoxitin | HGB | 10% | 6 | 2 | 7 | 0 | 0 | 0 |
| Stanford | cefoxitin | HGB | 25% | 11 | 1 | 2 | 1 | 0 | 0 |
| Stanford | cefoxitin | HGB | 50% | 9 | 4 | 2 | 0 | 0 | 0 |
| Stanford | cefoxitin | HGB | 100% | 3 | 1 | 0 | 1 | 0 | 0 |
| Stanford | cefoxitin | LR | 10% | 6 | 5 | 3 | 0 | 0 | 1 |
| Stanford | cefoxitin | LR | 25% | 13 | 0 | 2 | 0 | 0 | 0 |
| Stanford | cefoxitin | LR | 50% | 11 | 1 | 3 | 0 | 0 | 0 |
| Stanford | cefoxitin | LR | 100% | 3 | 1 | 1 | 0 | 0 | 0 |
| Stanford | ceftazidime | HGB | 10% | 6 | 3 | 2 | 1 | 0 | 3 |
| Stanford | ceftazidime | HGB | 25% | 6 | 4 | 2 | 0 | 3 | 0 |
| Stanford | ceftazidime | HGB | 50% | 5 | 3 | 4 | 2 | 1 | 0 |
| Stanford | ceftazidime | HGB | 100% | 2 | 2 | 1 | 0 | 0 | 0 |
| Stanford | ceftazidime | LR | 10% | 7 | 1 | 2 | 0 | 1 | 4 |
| Stanford | ceftazidime | LR | 25% | 8 | 4 | 1 | 2 | 0 | 0 |
| Stanford | ceftazidime | LR | 50% | 7 | 5 | 2 | 1 | 0 | 0 |
| Stanford | ceftazidime | LR | 100% | 2 | 1 | 2 | 0 | 0 | 0 |
| UTSW | cefepime | HGB | 10% | 7 | 3 | 1 | 3 | 0 | 0 |
| UTSW | cefepime | HGB | 25% | 9 | 2 | 3 | 1 | 0 | 0 |
| UTSW | cefepime | HGB | 50% | 9 | 3 | 1 | 2 | 0 | 0 |
| UTSW | cefepime | HGB | 100% | 3 | 1 | 1 | 0 | 0 | 0 |
| UTSW | cefepime | LR | 10% | 9 | 1 | 1 | 3 | 0 | 0 |
| UTSW | cefepime | LR | 25% | 9 | 2 | 3 | 1 | 0 | 0 |
| UTSW | cefepime | LR | 50% | 10 | 2 | 1 | 2 | 0 | 0 |
| UTSW | cefepime | LR | 100% | 4 | 0 | 1 | 0 | 0 | 0 |
| UTSW | ceftazidime | HGB | 10% | 12 | 3 | 0 | 0 | 0 | 0 |
| UTSW | ceftazidime | HGB | 25% | 15 | 0 | 0 | 0 | 0 | 0 |
| **Site** | **Drug** | **Model** | **Budget** | **α = 0** | **α = 0.05** | **α = 0.1** | **α = 0.25** | **α = 0.5** | **α = 1** |
| UTSW | ceftazidime | HGB | 50% | 15 | 0 | 0 | 0 | 0 | 0 |
| UTSW | ceftazidime | HGB | 100% | 5 | 0 | 0 | 0 | 0 | 0 |
| UTSW | ceftazidime | LR | 10% | 11 | 4 | 0 | 0 | 0 | 0 |
| UTSW | ceftazidime | LR | 25% | 15 | 0 | 0 | 0 | 0 | 0 |
| UTSW | ceftazidime | LR | 50% | 15 | 0 | 0 | 0 | 0 | 0 |
| UTSW | ceftazidime | LR | 100% | 5 | 0 | 0 | 0 | 0 | 0 |

